# The genetics of TDP43-Type-C neurodegeneration: a whole genome sequencing study

**DOI:** 10.1101/2025.01.25.25320561

**Authors:** Malik Nassan, Ivan Alejandro Ayala, Jennifer Sloan, Anna Bonfitto, Bobbi Stark, Serena Song, Marcus Naymik, Changiz Geula, Tamar Gefen, Elena Barbieri, Ignazio S. Piras, M-Marsel Mesulam, Matt J. Huentelman

## Abstract

Frontotemporal lobar degeneration-TDP Type C (TDP-C) is a unique neurodegenerative disease that starts by attacking the anterior temporal lobe leading to language and/or behavioral syndromes. Current literature on the genetic associations of TDP-C, which we have reviewed here, is uneven and lacks a discernible corpus of robust findings. In our study, we completed genome wide hypothesis-free analyses utilizing artificial Intelligence (AI) to identify rare and common variants associated with TDP-C. We then investigated *ANXA11* and *TARDBP* in a hypothesis-driven analysis, since it was recently shown that TDP-43 and Annexin A11 co-aggregate in all TDP-C cases.

1) Whole genome sequencing was completed to identify pathogenic rare variants prioritized with Illumina’s AI-based Emedgene software on 37 confirmed or probable TDP-C cases from the Northwestern-University Cohort. 2) A genome wide association study was then completed to identify common variants associated with TDP-C cases vs 290 controls. 3) Next, common and rare variants in *TARDBP, and ANXA11* were investigated in TDP-C vs controls.

These analyses identified novel genetic associations between *FIG4*, *UBQLN2*, *INPP5A*, and *ANXA11* with TDP-C. Of these *FIG4, UBQLN2 and ANXA11* have been associated previously with Amyotrophic lateral sclerosis (ALS). To further assess the observed potential genetic overlap between ALS and TDP-C, we leveraged Mendelian randomization (MR) to assess if the ALS genetic load is associated with TDP-C risk, and found evidence supporting this association.

The genetic association of *ANXA11* with TDP-C is particularly interesting in view of the recently discovered role of *Annexin A11* in forming heterodimers with TDP-43 in all abnormal precipitates, a feature not found in TDP-A or TDP-B, which have no similar predilection for the anterior temporal lobe. In addition to the observed overlap between ALS genetics/ genetic load and TDP-C, it is worth mentioning that *FIG4, INPP5A and ANXA11* have been implicated in the inositol metabolism pathway, a feature that remains to be elucidated mechanistically. Our TDP-C genetic literature review identified a surprising paucity of neuropathologically confirmed cases in published investigations. Nonetheless, the literature offers support for some of our findings and reemphasizes the absence of dominant or major pathogenic genes for TDP-C, another feature that sets this neuropathologic entity apart from TDP-A and TDP-B.

## Introduction

Neurodegenerative disorders are a major cause of morbidity and mortality with no available treatments that stop the disease progression, which is partly due to the limited understanding of the underlying pathogenesis.^1,2^ Frontotemporal lobar degeneration Type C (TDP-C) is a neurodegenerative disease that consistently starts by attacking the anterior temporal lobe (ATL).^3^ The resultant syndromes depend on the hemispheric asymmetry of atrophy, in which typically left ATL variant leads to semantic variant primary progressive aphasia (svPPA) while right ATL variant leads to associative agnosia or behavioral variant FTD syndrome (bvFTD).^4–8^ TDP-A and TDP-B neurodegenerative syndromes although associated with the same proteinopathy differ from TDP-C in which they do not have a consistent predilection for ATL and have nuclear and cytoplasmic preponderance of inclusions while TDP-C is characterized by long neurites concentrated mostly in upper layers of cortex.^4–6,9^

TDP-43 (Transactive response DNA binding protein of 43 kDa) is an intranuclear protein encoded by the *TARDBP* gene that is involved in RNA splicing, stabilization, and regulation of genes expression.^9,10^ The phosphorylated form of TDP-43 is a hallmark of amyotrophic lateral sclerosis (ALS) and a subset of frontotemporal dementia (FTD) disorders.^10^ Most recently, annexin A11 (encoded by *ANXA11* gene) was found to co-assemble with TDP-43 in aggregates in all TDP-C cases but much more rarely, if at all, in TDP-A or B.^11,12^ The unique characteristics of TDP-C makes this disease a promising target for disease-modifying interventions once its pathogenesis is identified.

Arguably, one of the first steps to understand this disease pathogenesis is through identifying its associated genes which can shed light to the implicated biological pathways and potential pharmacological targets. TDP-C is the least studied neurodegenerative syndrome from the molecular perspective, in which no known pathogenic genetic variants have been robustly identified.^13–15^ Current evidence suggests that both rare and common genetic variants may play a role in TDP-C risk. Some familial presumptive TDP-C cases have been reported,^16^ with existing evidence suggesting rare genetic variants are likely to play a role in TDP-C risk.^17^ Although common genetic variants have been proposed to contribute to the disease pathogenesis ^18^ the small sample size (even in international consortia) of this rare disease has been a limitation against identifying robust common variants.

Given the paucity of this literature and its rather meager content of pathologically proven TDP-C cases, we review the reported genetic associations in Table 1 in order to provide context for our investigations. The prior studies were limited either by lack of pathological confirmation of the diagnosis (for example, none of the reported *TARDBP* associations were based on pathologically confirmed TDP-C). Most - if not all - studies were limited by reduced power for traditional whole genome search strategies. In addition, the unique co-aggregation of TDP-43 and annexin A11 has not been known to previous genetic work and thus hypothesis driven studies did not focus on *ANXA11*.

In this study, we leveraged a well characterized (clinically and pathologically) cohort of TDP-C from Northwestern University.^5^ Using blood samples from pathologically confirmed or probable TDP-C cases and utilizing whole genome sequencing we ran two sets of analyses. We first completed genome wide hypothesis free analyses to identify rare variants (supported by Artificial Intelligence (AI)) and common variants associated with TDP-C. Then we completed hypothesis driven analyses focusing on TARDBP and ANXA11 genes, knowing the recent discovery of their co-assembly specifically in TDP-C cases.

## Materials and methods

### A- Review of the literature

We created Table 1 to review the genetic associations reported in the literature with TDP-C. We only reported associations specific to TDP-C and did not include associations with FTLD-TDP in general (knowing the different clinical and neuropathological characteristics between TDP-C and another FTLD-TDP pathologies and to reduce heterogeneity). We included large cohorts, family studies and case reports. We explained if the pathology was confirmed by autopsy or not. We also added our findings at the end of the table for the reader to be able to compare them to other studies.

### B- Whole genome sequencing study in the Northwestern TDP-C cohort

#### Participants

All participants have provided written consents to enroll in the cohort and for their samples to be used in genetic studies. This study was approved by Northwestern University Institutional Review Board.

Blood samples from TDP-C cases (n=37) were included in this study, which were selected from the clinically and pathologically well-characterized PPA cohort of the Mesulam Center at Northwestern University. In brief, 22 cases had confirmed TDP-C pathology and 15 cases were probable TDP-C cases [characterized with a clinical syndrome most likely – with about 90% correlation - to be due to TDP43-C (svPPA/ semantic dementia (SD), temporal pole as the most predominant region of atrophy, and when available biomarkers not consistent with Alzheimer’s disease)].^5^ Clinical diagnosis was based on the clinical evaluation of a single behavioral neurologist with expertise in PPA (M-Marsel Mesulam), using criteria described elsewhere.^6^ In brief, the svPPA diagnosis is based on prominent impairment of single word comprehension, and severe anomia (reflecting word comprehension as well as retrieval failures). Grammar and repetition are typically preserved, while speech can be vague and contains usually circumlocutions and semantic paraphasia.^6^ MRI of enrolled svPPA patients showed peak atrophy in dominant anterior temporal pole.^6^ Demographic and clinical characteristics for the included cases are summarized in table 2. The pathological characterization of the first 10 of the autopsied cases has been discussed elsewhere (for review please refer to Kawles A., et al).^9^

### Whole genome sequencing

WGS was performed using PCR-free library preps from Illumina to a cohort average coverage of 35X. Sequencing was performed using paired-end 151bp chemistry on the Illumina NovaSeq-X. Alignment and preprocessing were conducted using NVIDIA parabricks germline pipeline v4.1.0. The workflow includes alignment with BWA-mem, BAM sorting, marking duplicates, and base-quality score recalibration (BQSR). gVCFs were combined for variant joint call using *GLnexus* v1.2.6.

### WGS analyses to identify rare pathogenic variants

Variants were prioritized with Illumina’s Emedgene AI-based genomic analysis and interpretation platform (Illumina; https://emedgene.com; 2022), and individually inspected using Integrated Genome Viewer (IGV) browser to verify their quality. *Emedgene* is an Artificial Intelligence (AI) model and interpretation platform. *Emedgene* generates a shortlist of potential causative variants along with supporting evidence from the literature and curated databases, significantly reducing the time to interpret a case. The automated interpretation machine learning algorithms have been trained on thousands of patient samples and incorporate dozens of features typically used by geneticists to interpret cases. The algorithms use a proprietary knowledge graph of variants, genes, diseases, phenotypes, and their connections, which include information extracted from literature with natural language processing. Overall, the system automates time-intensive aspects of variant analysis and variant research and enhances lab analysis capabilities. The cases were analyzed with the AI tier 2 setting, which divides the high score variants into four main categories: Most Likely (ML), Most Likely genes with unknown significance (GUS) GUS, Candidates, and Candidate GUS, with ML variants having higher scores than Candidate Variants. The algorithm also divides variants that are in genes with a known disease connection and within GUS and restricts the ML list to the most likely variants.

### In-silico functional characterization of the identified likely pathogenic variants

Protein structures were downloaded from AlphaFold Protein Structure Database in Protein Data Bank (PDB) format.^19^ The target residue was changed using *PyMOL v3.0*, and the protein stability in both the wild-type and mutated protein was estimated using *Foldx v4*.^20^ Specifically, the protein stability refers to the Gibbs free energy change (ΔG) associated with the protein folding process, which measure how energetically favorable or unfavorable the folded state of the protein is compared to the unfolded state.

### GWAS analyses searching for common variants associated with TDP-C vs controls

We ran a GWAS on the 37 TDP-C cases vs 290 controls. The controls were taken from two different studies: the Banner Brain and Body Donation Program^21^ and the WGS Harmonization study from AMP-AD (accession number: syn22264775). In both cases, the controls included donors with no known dementia syndrome (lacking clinical and neuropathological criteria for AD diagnosis). Details about the control cohort are shown in supplemental Table S1. After alignment and preprocessing (see above), VCF files were filtered to exclude variants with depth < 10, genotype quality (GQ) < 20, and quality score (QUAL) < 30. INDELs and non-biallelic variants were removed, as well as SNPs located within 10 bp of an INDEL. Genotypes were called as heterozygous when the allelic balance was between 0.2 and 0.8. Additional samples quality controls included removal of sex mismatches, duplicated samples, samples with extreme heterozygosity (± 3 standard deviations from the average), and outliers. After quality control, the final sample size was 37 TDP-C vs. 242 CTL for a total of 5,067,777 variants with a minimum allele frequency larger than 5%. The number of controls decreased because of the presence of the duplicated samples identified using genetic data through the KING-robust relatedness estimator.^22^ The Genome-Wide Association study was conducted employing a mixed linear model as implemented in GEMMA,^23^ adjusting for sex, the top principal component (PC) and incorporating the relatedness matrix estimated using GEMMA from the data. The number of PCs to include was determined by running the GWAS including a variable number of PCs from 1 to 10 and assessing the lambda inflation factor. Including sex and the top PC, we obtained the lowest value of inflation (l = 1.001). Variants were deemed as significant at the genome-wide level when p < 5.0E-08, and then annotated using ANNOVAR.

### Hypothesis driven analyses focusing on *TARDBP, and ANXA11*

Using the same GWAS results, we then attempted to investigate common and rare variants in genes that have sufficient pathological evidence in the literature for being likely pathogenic for TDP-C based on forming histopathological protein aggregates in TDP-C cases (TARDBP, ANXA11). We used ANNOVAR to annotate variants as upstream/downstream when at least 1-kb from the transcription start/end site. A total of 30 common *ANXA11* variants (MAF > 5%), pruned at linkage disequilibrium threshold of R^2^ 0.6 (PLINK option: *--indep-pairwise 50 5 0.6*), were included in the analysis. Total of 247 rare ANXA11 variants (MAF < 1%) were included in the analysis. A total of 2 common TARDBP variants (MAF > 5%), pruned at linkage disequilibrium threshold of R^2^ 0.6, were included in the analysis. A total of 60 rare TARDBP variants (MAF < 1%) were included in the analysis. P values were adjusted using the Benjamini and Hochberg method.

### Mendelian randomization analyses

To evaluate if the genetic load for ALS is associated with TDP-C risk, we leverage a two-sample mendelian randomization (MR) approach. We selected 16 SNPs associated at a genome wide level of significance with ALS in the largest ALS GWAS to date to be genetic proxy of ALS genetic load. We removed C9orf72 SNP (rs2453555) from the ALS genetic proxy since C9orf72is known to be associated with TDP43- type A or B, but not TDP-C.^24^ We then ran a two-sample MR analysis testing if ALS genetic load can be associated with TDP-C risk using the inverse-variance weighted (IVW) method.^25^ To assess the robustness of the findings, we then ran MR model-based sensitivity analyses, in addition to heterogeneity, pleiotropy and outliers analyses, and created scatter plot and leave-one-out plot.^25,26^ Genetic associations with exposure and outcome were harmonized by aligning beta coefficients to the same effect allele. The MR analyses were conducted using the R-studio v2024.04.1+748 and the TwoSampleMR v 0.4.26 package.

## Data availability

The GWAS summary statistics are openly available in Synapse at https://www.synapse.org/Synapse:syn64465917

## Results

A comprehensive review of the literature for genetic variants found to be associated with TDP-C is included in Table 1. This was completed to help compare our results with the existing literature and to guide our hypothesis driven analyses. We illustrated the reported previous associations based on whether the TDP-C diagnosis was pathologically confirmed as TDP-C or clinically diagnosed. Of interest, the TDP-C literature in multiple studies hinted to genes that have been also implicated or linked to ALS (*TARDBP, ANXA11, L3MBTL1*) [Table 1]. *TARDBP* and *ANXA11* have been implicated in TDP-C at the genetic and neuropathological levels which was the reason we focused on them in our cohort. Of note, none of the previously reported cases that had a germline genetic association with *TARDBP* were pathologically confirmed. Furthermore, the majority of the reported associations in the literature are preliminary and require further studies and validation, including the largest cohort of TDP-C cases to date, in which the reported common variant has not been replicated and the rare variants were present in a number of healthy controls.^27^

## 1. Hypothesis free analyses in the Northwestern TDP-C cohort

In our cohort, 37 confirmed and probable TDP43-C cases were included in the WGS analyses. The demographics and clinical characteristics of the cohort are summarized in Table 2.

### Rare variants analyses

Two cases (5% of total cases) were found to have a likely pathogenic rare genetic variant in *FIG4* and *UBQLN2* [Table 3, Supplemented figures 1,2], both of which previously implicated in ALS. Further bioinformatic functional analyses have been completed to predict the molecular pathways implicated by those two genes. The *FIG4* rare variant is a splice donor variant that likely leads to a truncated transcript. The UBQLN2 rare variant causes a moderate decrease in the stability of the mutated protein ((ΔG):-2.899 kcal/mol). Such moderate instability is likely to cause biologically significant changes to the involved protein functions.

**Figure 1:**
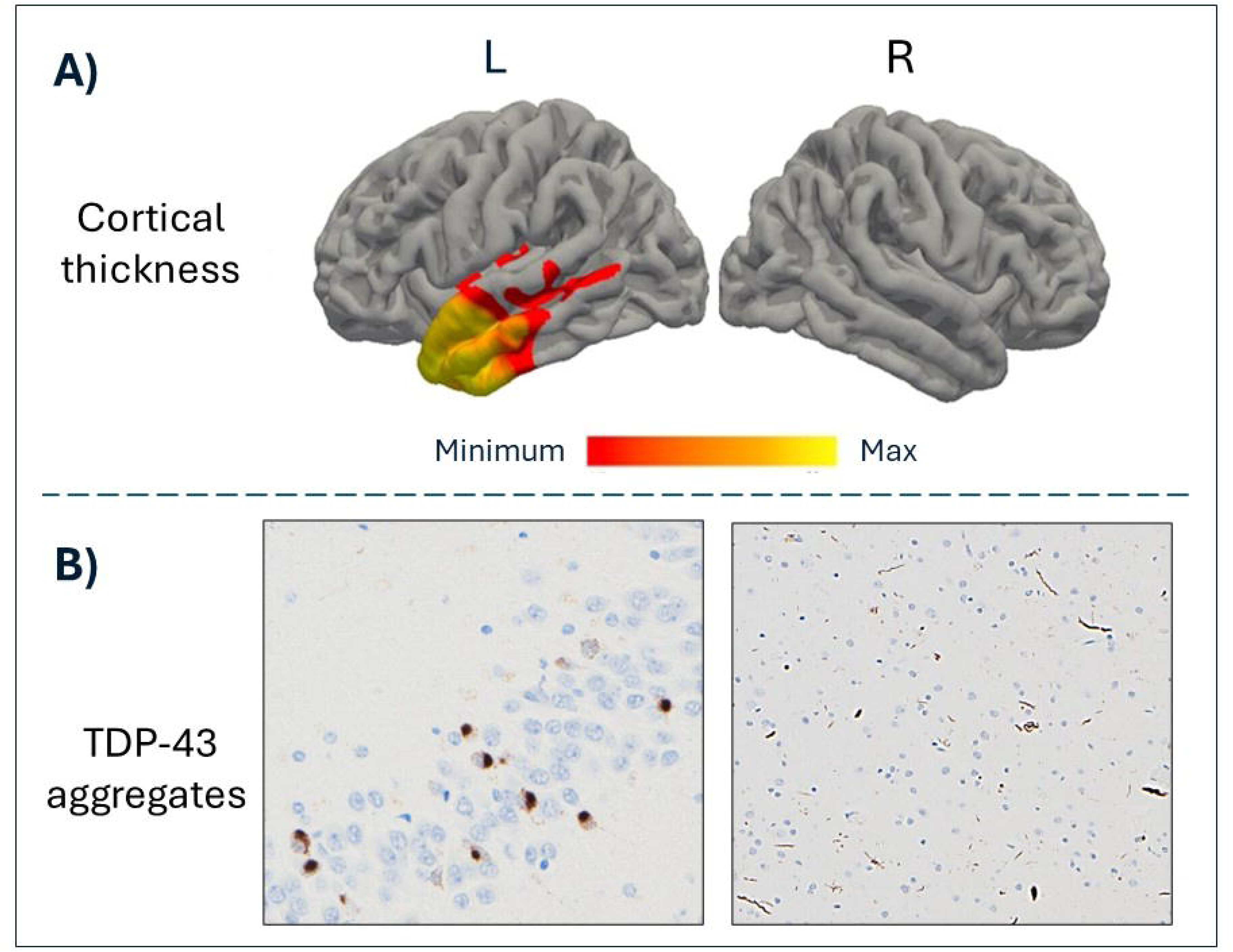
**A) Neuroimaging for the probable TDP-C case of svPPA with *FIG4* mutation**. These MRI based reconstructed images show predominant left anterior temporal lobe atrophy. **B) Pathological data for the confirmed TDP-C case with *UBQLN2* mutation.** Histopathological images showing TDP-43 aggregates of type C pattern.

We then sought to further evaluate those two cases from a clinical and neuropathological aspect. The first case with a variant in *FIG4* is a male patient a with svPPA presentation with predominant left temporal pole atrophy [Figure1-A], and negative clinical Amyloid PET scan. The svPPA diagnosis was based on expert assessment (M-Marsel Mesulam) and neuropsychological data showing that the most predominant cognitive impairments were in naming (using Boston Naming Test) and single word comprehension (using Peabody Picture Vocabulary Test-Form B). This patient had no signs of motor neuron disease. Family history was remarkable for a son formally diagnosed with a learning disability with a need for accommodations throughout school. The parents of this patient did not have dementia per report. The second case with *UBQLN2* variant is a female with svPPA syndrome and TDP-C confirmed pathology [Figure1-B]. There was also predominant left temporal pole atrophy at presentation, with a similar pattern to the one seen in Figure 1-A. There were no signs of motor neuron disease. The family history was positive for a brother with either learning or behavioral disability and was held back in school for a year. The parents of the patient did not have dementia per report.

### Common variants GWAS analysis

Using a GWAS approach, we found significant association between common variants in *INPP5A* with TDP-C vs controls (top associated SNP P= 1.26 x 10^-11^) [Table 3, Figure 2].

**Figure 2:**
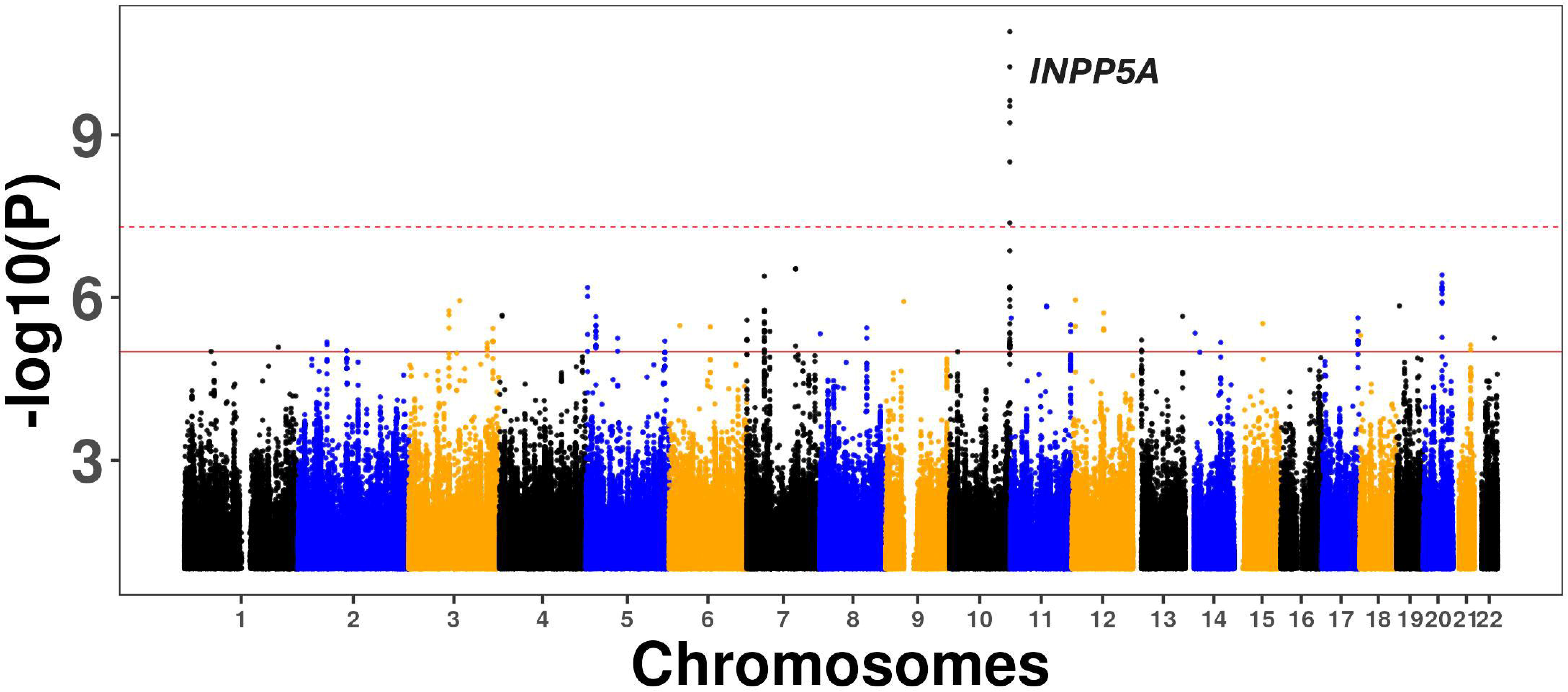
**GWAS Manhattan plot for TDP-C vs Controls**

## 2. Hypothesis driven analyses

When looking in our cohort at the common SNPs in the two genes (*TARDBP, ANXA11*) that have the most robust evidence from the literature and pathological studies to be associated with TDP-C, we found a significant association between the common variant rs113772135 in *ANXA11* gene (Beta regression coefficient = 0.23, Standard Error = 0.06, P=0.00036, BH adjusted P value= 0.012) [Table 5-A]. When looking in our cohort at the rare variants in the two genes (*TARDBP, ANXA11*), we found a significant association between the rare variant rs144114856 in *ANXA11* (BH adjusted P= 0.003) with TDP-C vs controls [Table 5-B] (complete results presented in Supplemented Tables S2,S3). No common or rare variants in *TARDBP* were found to be associated with TDP-C vs controls (BH adjusted P > 0.05) [supplemented Tables S4, S5].

The MR analyses included 11 SNPs from the ALS proxy that were available in the TDP-C vs control GWAS summary statistics [Supplemented Table S6]. We found a significant association between ALS genetic load and TDP-C risk, which was supported by sensitivity analyses (Table 6). MR scatter plot and leave-one-out plot support the robustness of the MR results and indicate lack of a single variant driving the association [Supplemented Figure S.3].

## Discussion

Although suggestive evidence exists, our literature review showed no robust genetic associations with TDP-C neurodegeneration. To date, there are no known major causative genetic mutations for TDP-C. In our cohort and using comprehensive WGS and AI approaches, we identified two novel rare genetic variants that are likely to contribute to the pathogenesis of TDP-C. Our results support the hypothesis that TDP-C can be caused by rare genetic variants even in the absence of family history, which could be explained by *de novo* mutations or mutations with low penetrance. Another potential explanation is the presence of an interaction with other genetic factors that can increase or decrease the risk of developing the TDP-C phenotype among carriers. One such example is the TMEM106B protective variant that protect GRN carriers from developing the FTD phenotype,^28^ or the Christchurch variant that protects APOE4 carriers from developing the AD phenotypes.^29^ Thus, the family members of genetic TDP-C cases could carry the risk variant but not have the phenotype due to protective genetic variants for example. On the other hand, environmental or epigenetic factors might contribute to the development of the TDP-C phenotype among the genetic variant carriers in a double hit model, in which the genetic variant is necessary but not sufficient to develop the disease. This proof-of-concept work supports further research using WGS to identify rare pathogenic variants in TDP-C patients even with the lack of family history.

Notably, both identified rare genetic variants in our cohort are in genes that have been associated with ALS, which is also a TDP43 neurodegenerative disease in the vast majority of cases. *FIG4* encodes a phosphoinositide 5-phosphatase which is implicated in trafficking of endosomal vesicles to the trans-Golgi network.^30^ Loss of functions of *FIG4* in mice results in degeneration of neurons.^30^ Compound heterozygotes mutations in *FIG4* in humans (indicating severe loss of function) leads to the recessively inherited severe form of Charcot-Marie-Tooth disease with early onset and involvement of both sensory and motor neurons.^30^ On the other hand, heterozygotes autosomal dominant nonsynonymous variants of *FIG4* (with less severe loss of function phenotype) have been associated with ALS and primary lateral sclerosis. ^30^ Our reported variant in *FIG4* linked to TDP-C here is a heterozygous genotype. A homozygous genotype of the same mutation has been reported in an autosomal recessive form for child onset leukoencephalopathy due to demyelination.^31^ Our finding is in line with literature that homozygous mutations in *FIG4* lead to early onset and severe neurologic disease, while a heterozygous mutation leads to a late onset neurodegenerative disease. One example of this phenomena can be seen among *GRN* mutation carriers, in which heterozygous carriers have late onset neurodegenerative FTD phenotype, while homozygous carriers have severe early onset neuronal ceroid lipofuscinosis-11 syndrome.^32,33^

*UBQLN2* is an intron-less gene encoding Ubiquilin-2 protein which is a member of the ubiquitin-like protein family (ubiquilins). In humans, there are four ubiquilin genes.^34^ Ubiquilins promote protein degradation through the ubiquitin-proteasome system and macroautophagy.^34,35^ *UBQLN2* mutations can lead to loss of function and have been found to impair misfolded proteins degradation and clearance pathways.^34,36–38^ In mouse models, pathogenic Ubqln2 was also found to cause neuron death through a toxic gain of function properties.^39^ In a familial ALS cohort, *UBQLN2* has been causally associated with ALS and FTD-MND (with behavioral executive dysfunction presentation) in an x-linked manner (no male-to-male transmission).^34^ ALS and FTD-MND patients (with or without *UBQLN2* mutation) have been found ubiquilin2 brain inclusions.^34^ In a subsequent genetic screening study of patients with non-familial ALS or FTD, 3 novel *UBQLN2* mutations were reported including a patient with pure bvFTD without MND.^40^

Our GWAS analysis provided evidence for the association between common variants in *INPP5A* in participants with TDP-C vs controls. *INPP5A* encodes a membrane-associated 43 kDa type I inositol 1,4,5-trisphosphate (InsP3) 5-phosphatase, which has been suggested to be implicated in dendritic spine morphogenesis in neurons.^41^ *INPP5A* plays a role in calcium signaling and the control of lipid exchange at membrane contact sites between cellular organelles and thus implicated in vesicular trafficking involved in the exchange of proteins, lipids, and metabolites.^42^ *INPP5A* has been implicated in spinocerebellar ataxia (SCA), in which *INPP5A* was downregulated in the cerebellum of SCA type 17 mice model.^43,44^ A microduplication in the 10q26.3 region (including part of *INPP5A* gene) was found in a patient with both childhood onset schizophrenia and Autism as well as her mother with schizophrenia, but not in the patient’s healthy sister.^41^ Intriguingly, in an epigenome-wide meta-analyses of blood-based DNA methylation levels for seven measures of cognitive functioning using data from 11 cohorts, cg12507869 in *INPP5A* was associated with phonemic verbal fluency (P = 2.5 × 10^-9^).^45^ This methylation site has a moderate correlation with brain methylation in Brodmann area 20 (ventral temporal cortex).^45^ In the same study, a methylation quantitative trait loci(QTL) analysis showed that rs113565688 was a cis methylation QTL for cg12507869.^45^ In another study, epigenome-wide association analysis on sorted neuronal and glia nuclei from postmortem human brain tissues (63 frontal and 65 temporal cortex samples derived from 52 healthy controls and 76 AD donors) showed differential methylation dynamics in *INPP5A* to be associated with aging in glia in controls. A limitation of this GWAS study is the limited sample size, as the results might suffer from sampling effects. However, because TDP-C is a very rare disease, obtaining samples for a large scale GWAS is not easily attainable.

Our hypothesis driven analyses showed supportive evidence for the association of *ANXA11* gene with TDP-C. Recently annexin A11 (encoded by *ANXA11* gene) was found to co-assemble with TDP-43 in aggregates in all TDP-C cases but much more rarely, if at all, in TDP-A or B.^11^ We sought to test the association of common and rare genetic variants in *ANXA11* with TDP-C. As listed in Table 1, two cases of TDP-C have been previously associated with likely pathogenic rare variants in *ANXA11*. Our findings provide evidence that in addition to rare variants, common variants in *ANXA11* might also be associated with TDP-C risk. The lack of association between *TARDBP* and TDP-C in our cohort could be due speculatively to the notion that *ANXA11* is the main driver of the neuropathology in TDP-C and that the accumulation of TDP-43 is a secondary consequence in the disease cascade. However, we acknowledge that the effect sizes of these variants, located in candidate genes, are limited if considered in a genome-wide context. These preliminary findings of the *ANXA11* variants association with TDP-C warrant further functional studies to establish the functional impact of the genetic variations in this gene.

From a mechanistic standpoint, it is of interest that we found an association between TDP-C with a rare (in *FIG4*) and common variants (in *INPP5A*) in genes that are part of the inositol metabolism pathway. Dysfunction in this pathway has been linked with excessive and toxic levels of intracellular Ca^2+^ that may contribute to the neurodegeneration process.^46–48^ ANXA11 is one of the annexins groups, which are Ca2+-regulated phospholipid-binding proteins that are implicated in the cell life cycle, apoptosis and exocytosis.^49^ *ANXA11* encodes a protein that leads RNA granules to lysosomes as they traffic in axons, with an N-terminus that contains a low-complexity domain, and C-terminus with calcium-binding domains.^50^ ANXA11 has been shown to bind to phosphatidylinositol 3-phosphate (PI(3)P)-containing giant unilamellar vesicles (composed of lysosomal membrane lipids) in a Ca2+-dependent manner and is dependent on the structured C-terminal annexin repeat domain.^51,52^ Taken together, our findings suggests that TDP-C neurodegeneration might emerge due to dysregulation in the inositol-calcium pathway leading to RNA to lysosomes trafficking abnormalities.

Our post-hoc analyses provided further supportive evidence of the association between ALS genes with TDP-C. The MR analyses showed robust association between ALS genetic load and risk for TDP-C, as supported by multiple lines of sensitivity analyses. Despite the relatively small sample size, those analyses further support that the accumulation of common variants associated with TDP-43 pathology (in this case ALS common variants) might be associated with increased risk for TDP-C neurodegeneration.

In this cohort, we reported novel rare genetic variants (in *FIG4* and *UBQLN2*) and common variants (in *INPP5A)* that are associated with TDP-C neurodegeneration. We also reported novel common and rare variants in *ANXA11* that are associated with TDP-C. Our overall results point to the possible overlap between ALS genes and TDP-C and implicate the inositol-calcium pathway as an underpinning mechanism for the RNA to lysosomes trafficking abnormalities in TDP-C neurodegeneration. Further functional studies of the identified genes could further illuminate the pathophysiological mechanism of this enigmatic disease and pave the way for targeted and personalized interventions. In addition, specialized cognitive genetic clinics may consider ALS genetic panel or WGS in suspected TDP-C patients even in the absence of family history. However, genetic counseling and a discussion of benefit vs risk and cost effectiveness and the actionability of the discovered genetic data should take place in the clinical setting as indicated. Our data support further studies of both rare and common genetic variants as risk factors for TDP-C neurodegeneration.

## Data Availability

Data can be accessed though the listed database in the manuscript.

## Acknowledgements

The results published here are in whole or in part based on data obtained from the AD Knowledge Portal (https://adknowledgeportal.org). The ROSMAP study data were provided by the Rush Alzheimer’s Disease Center, Rush University Medical Center, Chicago. Data collection was supported through funding by NIA grants P30AG10161 (ROS), R01AG15819 (ROSMAP; genomics and RNAseq), R01AG17917 (MAP), R01AG30146, R01AG36042 (5hC methylation, ATACseq), RC2AG036547 (H3K9Ac), R01AG36836 (RNAseq), R01AG48015 (monocyte RNAseq) RF1AG57473 (single nucleus RNAseq), U01AG32984 (genomic and whole exome sequencing), U01AG46152 (ROSMAP AMP-AD, targeted proteomics), U01AG46161(TMT proteomics), U01AG61356 (whole genome sequencing, targeted proteomics, ROSMAP AMP-AD), the Illinois Department of Public Health (ROSMAP), and the Translational Genomics Research Institute (genomic). Additional phenotypic data can be requested at www.radc.rush.edu. The Mount Sinai Brain Bank data were generated from postmortem brain tissue collected through the Mount Sinai VA Medical Center Brain Bank and were provided by Dr. Eric Schadt from Mount Sinai School of Medicine. The Mayo RNAseq study data was led by Dr. Nilüfer Ertekin-Taner, Mayo Clinic, Jacksonville, FL as part of the multi-PI U01 AG046139 (MPIs Golde, Ertekin-Taner, Younkin, Price). Samples were provided from the following sources: The Mayo Clinic Brain Bank. Data collection was supported through funding by NIA grants P50 AG016574, R01 AG032990, U01 AG046139, R01 AG018023, U01 AG006576, U01 AG006786, R01 AG025711, R01 AG017216, R01 AG003949, NINDS grant R01 NS080820, CurePSP Foundation, and support from Mayo Foundation. Study data includes samples collected through the Sun Health Research Institute Brain and Body Donation Program of Sun City, Arizona. The Brain and Body Donation Program is supported by the National Institute of Neurological Disorders and Stroke (U24 NS072026 National Brain and Tissue Resource for Parkinson’s Disease and Related Disorders), the National Institute on Aging (P30 AG19610 Arizona Alzheimer’s Disease Core Center), the Arizona Department of Health Services (contract 211002, Arizona Alzheimer’s Research Center), the Arizona Biomedical Research Commission (contracts 4001, 0011, 05-901 and 1001 to the Arizona Parkinson’s Disease Consortium) and the Michael J. Fox Foundation for Parkinson Research.

## Supplementary material

Supplementary material is available at *Brain Communications* online.

