## Supplementary material for "The genetics of TDP43-Type-C neurodegeneration: a whole genome sequencing study": Tables

**Table1:** review of the literature of previously implicated genes in confirmed and probable TDP-C neurodegeneration.

| **Genes** | **Approach** | **Study** |
| --- | --- | --- |
| *C19orf52* (common variant)  *RBPJL* (rare variant)  *L3MBTL1* (rare variant) | WGS of 467 pathologically confirmed TDP-C cases and 3,153 Controls, results not replicated | Pottier. C., et al., Deciphering Distinct Genetic Risk Factors for FTLD-TDP Pathological Subtypes via Whole-Genome Sequencing. *medRxiv,* 2024 |
| Rare variants in *TARDBP* and *C9orf72* | Candidate gene study of 122 svPPA/SD cases, no pathological diagnosis | Ramos EM, et al. Genetic screen in a large series of patients with primary progressive aphasia. Alzheimers Dement. 2019 |
| 791 differentially expressed genes in the FTLD-TDP-C group compared to controls | bulk RNA-sequencing on the frontal cortex from pathologically confirmed TDP-C Cases (n= 22) and Controls (n= 24) | Pottier. C., et al, Shared brain transcriptomic signature in TDP-43 type A FTLD patients with or without GRN mutations, *Brain*, 2022 |
| *HNF4A, NR5A1, TAL1, SLC2A4, PSEN1, KRT81, MYBL2, UBE2I, EBI3, BATF, ARFRP1, NR6A1, PACS1, PELP1, and TEF* | Bio-informatic search for common genetic variants supported by gene expression data. Cases in first stage: publicly available 361 with semantic dementia vs 4308 controls.  Cases in second stage: publicly available dataset FTLD with ubiquitinated inclusions (n=10) vs controls (n=11)* | Bonham, L.W., *et al.* Genetic variation across RNA metabolism and cell death gene networks is implicated in the semantic variant of primary progressive aphasia. *Sci Rep,* 2019. |
| *ANXA11* (rare variant) | Candidate gene study. One out 34 pathologically confirmed TDP-C cases had possible pathogenic ANXA11 rare variant | Robinson, J.L., et al. Annexin A11 aggregation in FTLD–TDP type C and related neurodegenerative disease proteinopathies. *Acta Neuropathol,* 2024. |
| *ANXA11* (rare variant) | Case report of anomia and prosopagnosia with predominant right-ATL syndrome, no pathological diagnosis | Kim, E.J., et al. Semantic variant primary progressive aphasia with a pathogenic variant p.Asp40Gly in the ANXA11 gene. *Eur J Neurol,* 2022 |
| *TARDBP* | Deep whole exome sequencing searching for Brain somatic mutation from the middle temporal gyrus, in which 2/16 cases with TDP-C confirmed pathology were found to have somatic TARDBP mutations. This study excluded germline variants | van Rooij, J., et al. Somatic TARDBP variants as a cause of semantic dementia, *Brain*, 2020, |
| *TARDBP* | Case series of FTD syndromes with TARDBP pathogenic mutation, 5 of 29 patients with TARDBP mutations have svPPA/SD, no pathological diagnosis | Caroppo. P., et al, Defining the spectrum of frontotemporal dementias associated with TARDBP mutations, *Neurology Genetics,* 2016 |
| *TARDBP* | Case report of a Patient with svPPA/SD and a sibling with ALS, no pathological diagnosis | González-Sánchez, M., et al. TARDBP mutation associated with semantic variant primary progressive aphasia, case report and review of the literature. *Neurocase*. 2018 |
| *TARDBP* | Case report of family with TARDBP mutation, in which Three members of the family had different age at onset and clinical presentations (one with ALS, one with SD, one with bvFTD-MND). They all carried c.1147A!G (p.I383V) missense mutation in the TARDBP gene coding sequence, no pathological diagnosis | Cheng. Y.W., et al. A single nucleotide TDP-43 mutation within a Taiwanese family: A multifaceted demon. *Amyotroph Lateral Scler Frontotemporal Degener*. 2016 |
| *TARDBP* | A cohort of 149 FTLD-MND cases and 400 controls screened for TARDBP mutations. One patient with svPPA/SD presentation followed by MND had TARDBP p.Gly295Ser mutation. One sibling with the same mutation had ALS presentation, father with ALS presentation and unknown genotype, no pathological diagnosis | Benajiba, L., et al. TARDBP mutations in motoneuron disease with frontotemporal lobar degeneration. *Ann Neuro*, 2009 |
| *TUBA4A* | Whole exome sequencing of a patient with SD diagnosis and family history of parkinsonism; pathological diagnosis confirmed TDP-C. Patient was found to have novel frameshift mutation in the TUBA4A gene leading to a decrease in total TUBA4A mRNA and protein levels. | Van Schoor, E., et al. Frontotemporal Lobar Degeneration Case with an N-Terminal TUBA4AMutation Exhibits Reduced TUBA4A Levels in the Brain and TDP-43 Pathology. *Biomolecules*, 2022 |
| *UBQLN2* (rare pathogenic variant)  *FIG4* (rare pathogenic variant)  INPP5A (common variants)  ANXA11(common and rare variants) | This study leveraged WGS on a cohort of autopsy confirmed (n=22) or probable (n=15) TDP-C cases with svPPA (total n=37). *UBQLN2, FIG4,* INPP5A were identified in hypothesis free genome wide analyses, while ANXA11 was associated with TDP-C based on hypothesis driven analysis. Mendelian randomization analysis showed association between ALS genetic load and TDP-C | The current study |

**Table 2:** The demographic and clinical characteristics of the TDP-C cohort

| **Cases** | **N** | **Females (%)** | **Mean Age onset (range)** | **Mean Age at death**  **(range)** | **APOE2 carriers (%) *** | **APOE4 carriers (%) *** | **Phenotypes other than svPPA** |
| --- | --- | --- | --- | --- | --- | --- | --- |
| Total | 37 | 20 (54%) | 59.3 (50-79) | NA | 5 (14%) | 5 (14%) | - |
| Autopsy confirmed | 22 | 14 (64%) | - | 72 (61-93) | - | - | 1 m-PPA,  1 SD,  1 bvFTD |
| Clinically diagnosed | 15 | 6 (40%) | - | NA | - | - | 1 SD,  1 isolated anomia |

*No cases with homozygous APOE2,2 or APOE 4,4, and no cases with APOE 2,4 genotype

**Table 3:** TDP-C cases with rare likely pathogenic variants in our cohort

| **Case** | **sex** | **Clinical diagnosis** | **Pathological diagnosis** | **APOE** | **Gene** | **Mutation** | **Genotype** | **allele frequency in gnomAD** | **Combined Annotation Dependent Depletion (CADD)+** | **Note** |
| --- | --- | --- | --- | --- | --- | --- | --- | --- | --- | --- |
| 1 | Male | svPPA | Probable TDP-C, with negative clinical Amyloid PET scan | 3,3 | FIG4 | NM_014845.6:c.2459+1G>A; splice donor variant (truncated) | heterozygous | 0.00002646 | 34 | ALS gene |
| 2 | Female | svPPA | Confirmed TDP-C | 2,3 | UBQLN2 | NM_013444.4:c.431G>A;  NP_038472.2 p.Gly144Glu | Heterozygous, missense mutation | 0.00001797 | 23.2 | ALS gene |

+ score > 10 is likely pathogenic

**Table 4.** GWAS common variants associated with TDP-C vs controls in our cohort at genome wide level of significance

| **avsnp151** | **OR** | **beta** | **se** | **p_wald** | **Func.refGene** | **Gene.refGene** |
| --- | --- | --- | --- | --- | --- | --- |
| rs35845236 | 1.45 | 0.37 | 0.05 | 1.26E-11 | intronic | INPP5A |
| rs61861422 | 1.36 | 0.30 | 0.04 | 5.61E-11 | intronic | INPP5A |
| rs61862807 | 1.35 | 0.30 | 0.06 | 2.35E-10 | intronic | INPP5A |
| rs35253505 | 1.42 | 0.35 | 0.05 | 2.99E-10 | intronic | INPP5A |
| rs12766156 | 1.45 | 0.37 | 0.06 | 6.01E-10 | intronic | INPP5A |
| rs34460796 | 1.33 | 0.29 | 0.05 | 3.17E-09 | intronic | INPP5A |
| rs745513405 | 1.21 | 0.19 | 0.03 | 4.22E-08 | ncRNA_intronic | LOC107984282 |

**Table 5:** hypothesis driven common and rare variants associated with *ANXA11* in our cohort

| **Rs ID** | **n_miss** | **allele1** | **allele0** | **Allele Frequency** | **OR** | **beta** | **se** | **p_wald** | **Func.refGene** |
| --- | --- | --- | --- | --- | --- | --- | --- | --- | --- |
| rs113772135 | 0 | T | C | 0.052 | 1.26 | 0.23 | 0.06 | 0.00036* | UTR3 |
| rs2789686 | 1 | T | C | 0.099 | 1.16 | 0.15 | 0.05 | 0.002 | UTR3 |
| rs1079242 | 2 | G | A | 0.498 | 0.94 | -0.06 | 0.03 | 0.03 | intronic |
| rs61860017 | 0 | A | T | 0.065 | 0.90 | -0.11 | 0.06 | 0.046 | intronic |
| rs34074920 | 2 | G | A | 0.096 | 1.07 | 0.07 | 0.05 | 0.11 | intronic |

A- Top common variants in *ANXA11* significantly associated with TDP-C vs controls

* Statistically significant after P value multiple testing correction= 0.012

B- Top rare variants in *ANXA11* significantly associated with TDP-C vs controls

| **Rs ID** | **n_miss** | **allele1** | **allele0** | **Allele Frequency** | **OR** | **beta** | **se** | **p_wald** | **Func.refGene** |
| --- | --- | --- | --- | --- | --- | --- | --- | --- | --- |
| rs144114856 | 0 | A | G | 0.009 | 1.95 | 0.67 | 0.15 | 9.43E-06* | intronic |
| rs184651165 | 0 | C | T | 0.005 | 1.73 | 0.55 | 0.20 | 0.005 | intronic |
| rs1180489738 | 7 | A | C | 0.002 | 2.53 | 0.93 | 0.34 | 0.006 | intronic |
| rs1262499074 | 0 | T | C | 0.002 | 2.46 | 0.90 | 0.34 | 0.008 | intronic |
| rs753662122 | 0 | A | G | 0.002 | 2.46 | 0.90 | 0.34 | 0.008 | intronic |

* Statistically significant after P value multiple testing correction = 0.003

**Table 6:** MR primary and sensitivity analyses testing the association between ALS genetic load and TDP-C in our cohort

| **MR method** | **n-snp** | **beta** | **se** | **P** |
| --- | --- | --- | --- | --- |
| Inverse-variance-weighted | 11 | 0.31 | 0.11 | 0.0046 |
| MR-Egger | 11 | 0.69 | 0.48 | 0.18 |
| Weighted-median | 11 | 0.40 | 0.14 | 0.005 |
| Weighted-mode | 11 | 0.41 | 0.18 | 0.04 |
| mr_heterogeneity Inverse variance weighted | | | | 0.54 ^#^ |
| mr_pleiotropy egger_intercept | | | | 0.44 * |
| Outliers testing MR-Global PRESSO | | | | 0.57 ^◊^ |

\# Indicating no heterogeneity

* Indicating no pleiotropy

◊ Indicating no outliers
