## supplement for "The genetics of TDP43-Type-C neurodegeneration: a whole genome sequencing study"

**Supplemental figures:**

**Figure S.1**: **Reads alignment of the region encompassing FIG4 mutation**

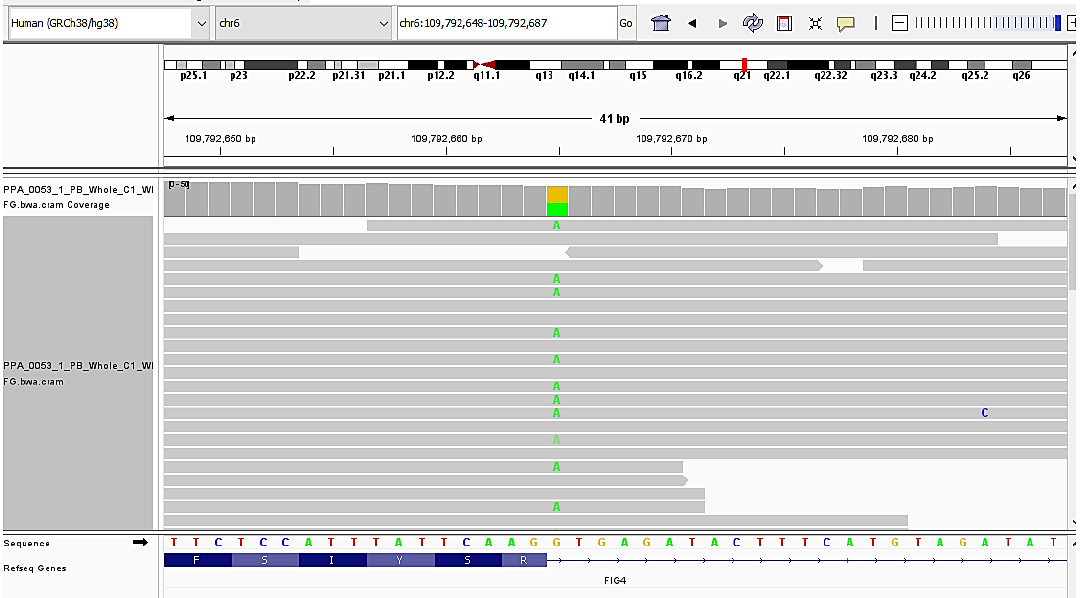

**Figure S.2:** **Reads alignment of the region encompassing *UBQLN2* mutation**

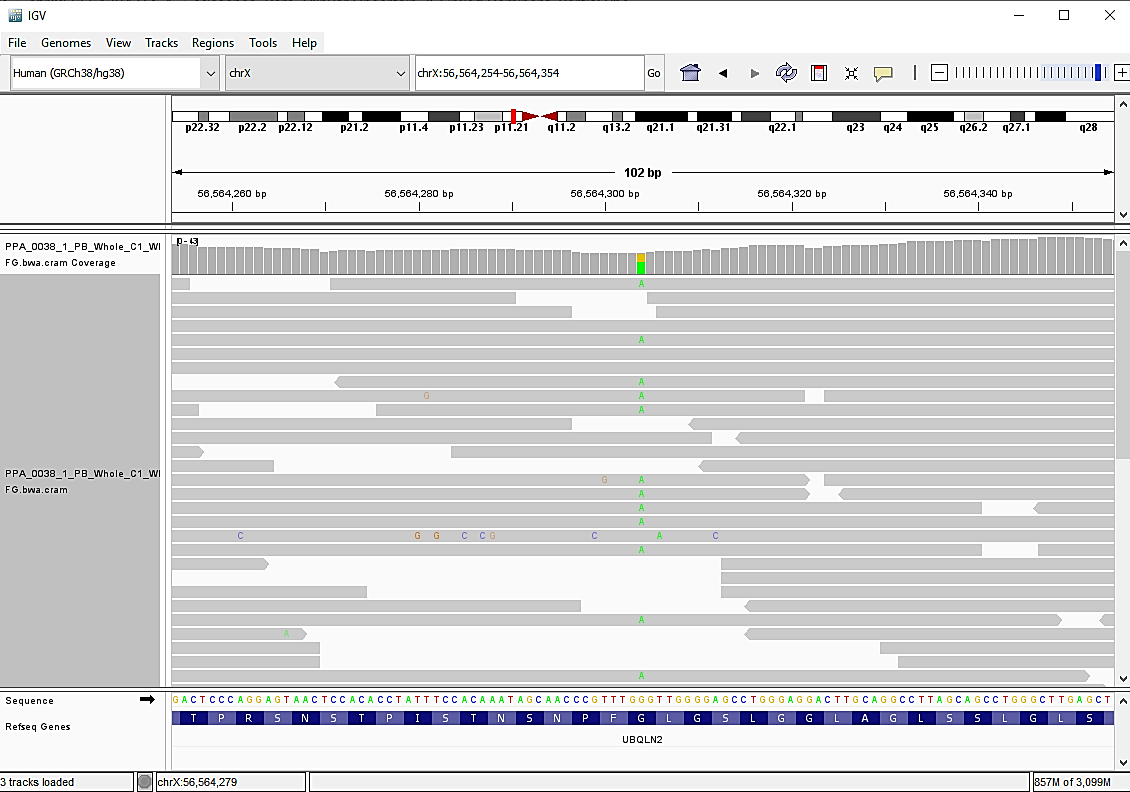

**Figure S.3: MR analyses.** A) MR scatter plot showing various MR models testing the association between ALS genetic load and TDP-C. B) leave-one-out figure testing whether an outlier was driving the association between ALS genetic load and TDP-C.

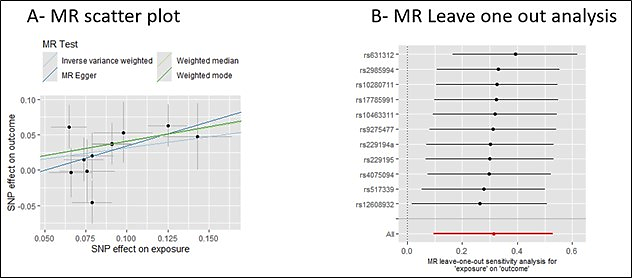
